# CORRELATION OF BODY COMPOSITION (BODY MASS INDEX, WAIST-TO-HIP RATIO, WAIST-TO-HEIGHT RATIO, NECK CIRCUMFERENCE) WITH FITNESS INDEX USING HARVARD STEP TEST

**DOI:** 10.1101/2023.07.13.23292635

**Authors:** Chinmoyee Baruah Hazra

## Abstract

**BACKGROUND:** Despite many clear benefits of active lifestyle lack of physical activity are common mostly among students, which further lead to alteration in body compositions. This alteration in body composition is the leading cause of various diseases as well as decrease in Fitness Index level.

**OBJECTIVE:** To correlate the body compositions i.e. Body Mass Index, Waist-to-Hip Ratio, Waist-to-Height Ratio, Neck Circumference with Fitness Index using Harvard Step Test among students of PEWS Group of Institutions and also to find which body composition is the strongest predictor of Fitness Index.

**METHODOLOGY:** 100 subjects were selected randomly in the study and were divided into two groups i.e Group A (Male) and Group B (Female), each having 50 subjects. For each subject BMI, WHR, WHtR, NC was measured. Harvard Step Test was performed and FI was calculated using it.

**RESULT:** Based on the statistical analysis i.e Karl Pearson’s Coefficient of Correlation, BMI, WHR, WHtR, NC all are negatively correlated with FI for both males and females. BMI, WHtR, NC of females and WHR of males have greater negative correlation with FI.

**CONCLUSION:** There is correlation between body compositions i.e. BMI, WHR, WHtR, NC and FI. BMI is the best predictor of FI in both males and females; also it has been observed that the males are fitter than the females.

## INTRODUCTION

Physical fitness is a general state of health and well-being and more specifically, the ability to perform. Before the industrial revolution, fitness was the capacity to carry out the daily activities without undue fatigue. However with automation and changes in lifestyles physical fitness is now considered a measure of the body’s ability to function efficiently and effectively in work and leisure activities, to be healthy to resist diseases and to meet emergency situations. Despite many clear benefits of active lifestyle lack of physical activities are common mostly among students^[1]^.

Body composition is an analysis of the percentage of stored fat in a body.Often clinicians choose simple body measurements to screen individuals for risk of disease associated with being overweight and obese: Body mass index (BMI), Waist circumference (WC), Waist-to-hip ratio (WHR)^[2]^.In recent years, two new indicators proposed to evaluate central adiposity Waist-to-height ratio (WHtR), Neck circumference (NC)^[3]^.

Cardiovascular disease is a leading cause of global mortality, accounting for almost 17 million deaths annually. The rate of cardiovascular disease is accelerating worldwide and one of the causes is the dramatic increase in the prevalence of obesity with its related complications of hypertension, hyperlipidemia, diabetes etc. The greater the obesity level, the body fatness or the abdominal obesity the greater the risk of developing cardiovascular disease. Other risk factor of cardiovascular disease include physical inactivity, elevated psychosocial stress and inappropriate diet^[4]^.

The Harvard Step Test is a cardiac stress test for detecting and diagnosing cardiovascular diseases. It is a good measurement of fitness and person’s ability to recover after strenuous exercise. The more quickly the heart rate returns to resting, the better shape the person is in. In this test, the subject takes steps up and down on a platform of height 50 cm or 20 inches in a cycle of one step per two seconds. The rate of 30 steps per minute must be held up for 5 min or until exhaustion. Exhaustion is the point at which the subject cannot maintain the stepping rate for 15 seconds. The subject immediately sits down on completion of the test, and the heartbeats are counted for 1 to 1.5, 2 to 2.5, 3 to 3.5 minutes ^[5]^. Now, the fitness index is measured using the formula:

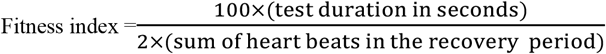

The outcomes of the equation are^[6]^:

### NEED OF STUDY

- There is lack of correlation studies regarding BMI, WHR, WHtR and NC with fitness index.
- There is lack of studies correlating these body compositions with Harvard Step Test in the age range of 18-35 years.

### OBJECTIVE OF THE STUDY

- The objective of the study is to correlate the body compositions i.e. Body Mass Index, Waist-to-Hip Ratio, Waist-to-Height Ratio, Neck Circumference with Fitness Index using Harvard Step Test among students of PEWS Group of Institutions and also to find which body composition is the strongest predictor of Fitness Index.

### HYPOTHESIS

#### NULL HYPOTHESIS

There will be no correlation between body compositions (Body Mass Index, Waist-to-Hip ratio, Waist-to-Height ratio, Neck Circumference) and Fitness Index using Harvard Step Test.

#### ALTERNATIVE HYPOTHESIS

There will be correlation between body compositions (Body Mass Index, Waist-to-Hip ratio, Waist-to-Height ratio, Neck Circumference) and Fitness Index using Harvard Step Test.

### RESEARCH QUESTION

- Is there any correlation between BMI & FI among females and males?
- Is there any correlation between WHR & FI among females and males?
- Is there any correlation between WHtR & FI among females and males?
- Is there any correlation between NC & FI among females and males?

### METHODOLOGY

The study was an exploratory study. Before selection of the subjects the study was explained and a written consent was taken from the subjects. 100 students out of which 50 males and 50 females of age group 18-35 years were randomly selected satisfying the inclusion criteria.

The subjects were taken from PEWS Group of Institutions, Bonda(Amgaon), Guwahati-26. This study was approved by the ethical committee of College of Physiotherapy and Medical Sciences; Bonda(Amgaon), Guwahati-26. Participants were asked not to take any heavy meal 2 hrs prior the test. Individuals having any cardio-respiratory compromise, severe neurological and musculoskeletal disorders limiting their mobility, individuals with psychiatric disorders with disruptive behavior, lack of cooperation and concentration were excluded from the study. Study duration was for 3 month.

### PROCEDURE

Participants were assigned into two groups:-

Group A: This group consists of 50 male students.

Group B: This group consists of 50 female students.

For both the groups same procedure was followed. Firstly BMI, WHR, WHtR and NC of students were measured and the procedure to perform the step test was demonstrated to them. They performed the step test one by one for 5 min or until and unless they were exhausted.

Immediately after completion of the test they were asked to sit down on a chair and their heartbeats were counted for 1 to 1.5 min, 2 to 2.5 min and 3 to 3.5 min. Now, their fitness index was calculated.

### BODY MASS INDEX (BMI)

The subjects were asked to stand erect and were asked to remove shoes and socks. Their height was recorded to nearest 0.1 cm and the weight was recorded to nearest 0.1 kg. Now, BMI was calculated using weight in kilograms divided by square of height in meters.

### WAIST-TO-HIP RATIO (WHR)

Waist circumference (in cm) was measured at the midpoint between the lower margin of last palpable rib and the top of the iliac crest and the Hip circumference (in cm) was measured around the widest portion of the buttocks using measuring tape. Now to calculate WHR, we divide waist circumference with hip circumference. WHR above 0.90 for males and above 0.85 for females is the indicator of health risk^[7]^.

### WAIST-TO-HEIGHT RATIO (WHtR)

The measurement of waist circumference (in cm) and the height (in cm), which was already recorded for BMI and WHR calculation. For WHtR we divided waist circumference with height. Males WHtR ranging from 0.43-0.52 and females 0.42-0.48 are categorized as healthy^[8,9]^.

### NECK CIRCUMFERENCE (NC)

Using measuring tape NC (in cm) was measured at a level of laryngeal prominence (Adam’s apple) in the midline formed by the thyroid cartilage at approximately C4 vertebra. Males with NC value of 38.5 cm and females with 34.5 cm are considered to be the optimal cutoffs for identifying visceral obesity^[10]^. And also cut off of 32.5 cm among females and 35.5 cm among males was best critical cut off to screen cardiovascular risk^[11]^.

### HARVARD STEP TEST

The subjects took the test steps up and down on platform of height 50 cm in a cycle of one step per two seconds or 30 steps per min for 5 min or until exhaustion. After completion the subject was asked to immediately sit on chair, and the heartbeats are counted for 1 to 1.5 min, 2 to 2.5 min and 3 to 3.5 min. Now, the fitness index is calculated using the formula :

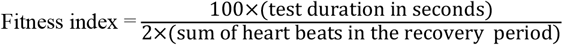

## DATA ANALYSIS AND RESULT

The collected data were analysed using SPSS Software version 20.0. Karl Pearson’s Coefficient of Correlation was performed to find the correlation of Body composition (Body mass index, Waist-to-hip ratio, Waist-to-height ratio, Neck circumference) with Fitness Index using Harvard Step Test among the students of PEWS Group of Institutions, Guwahati.

Karl Pearson’s Coefficient of Correlation is a statistical formula that measures the strength and relationship between two variables. It is denoted by ‘r’, and it ranges between -1 and +1. Nearer the value is to -1 or +1, greater is the correlation.

If the ‘r’ value is negative it denotes, with increase in one variable the other decreases or vice-versa. If the ‘r’ value is positive it denotes, with increase in one variable the other also increases or vice-versa. But if the ‘r’ value is 0 it denotes, there is no relation between the two variables.

### DATA ANALYSIS

#### Conclusion

From the table 3 we can conclude that BMI of females is more negatively correlated with FI than the males.

**Table 1:** Outcomes of Fitness index according to Harvard Step Test.

| **RATING** | **FITNESS INDEX** |
| --- | --- |
| Excellent | >96 |
| Good | 83-96 |
| Average | 68-82 |
| Low average | 54-67 |
| Poor | <54 |

**Table 2:** Classification of overweight and obesity by BMI, WC and Associated disease risk.

|  |  |  | **DISEASE RISK** |  |
| --- | --- | --- | --- | --- |
| Classification | BMI (Kg/m<sup>2</sup>) | Obesity class | Men≤ 40in.<br>(102 cm)<br>Women≤ 35in.<br>(88 cm) | Men>40in.<br>(102 cm)<br>Women>35in.<br>(88 cm) |
| Underweight | < 18.5 |  | - | - |
| Normal | 18.5-24.9 |  | - | - |
| Overweight | 25.0-29.9 |  | Increased | High |
| Obesity | 30.0-34.9 | I | High | Very high |
|  | 35.0-39.9 | II | Very high | Very high |
| Extreme obesity | ≥ 40 | III | Extremely high | Extremely high |

**Table 3:** Correlation of the BMI with the FI.

|  | 'r' value |
| --- | --- |
| MALE | -0.26552 |
| FEMALE | -0.36988 |

#### Conclusion

From the table 4 we can conclude that WHR of males is more negatively correlated with FI than the females.

**Table 4:** Correlation of WHR with the FI.

|  | 'r' value |
| --- | --- |
| MALE | -0.11709 |
| FEMALE | -0.11026 |

#### Conclusion

From the table 5 we can conclude that WHtR of females is more negatively correlated with FI than the males.

**Table 5:** Correlation of WHtR with the FI.

|  | 'r' value |
| --- | --- |
| MALE | -0.19596 |
| FEMALE | -0.29028 |

#### Conclusion

From the table 6 we can conclude that NC of females is more negatively correlated with FI than the males.

**Table 6:** Correlation of NC with the FI.

|  | 'r' value |
| --- | --- |
| MALE | -0.12941 |
| FEMALE | -0.32724 |

#### Conclusion

From the table 7 we see that the fitness index of males is more than the females.

**Table 7:** Mean of the FI of both males and females.

|  | MEAN |
| --- | --- |
| MALE | 58.782 |
| FEMALE | 47.552 |

#### Conclusion

From the table 8 we can conclude that for both males and females BMI is more negatively correlated with FI than any other body composition.

**Table 8:**
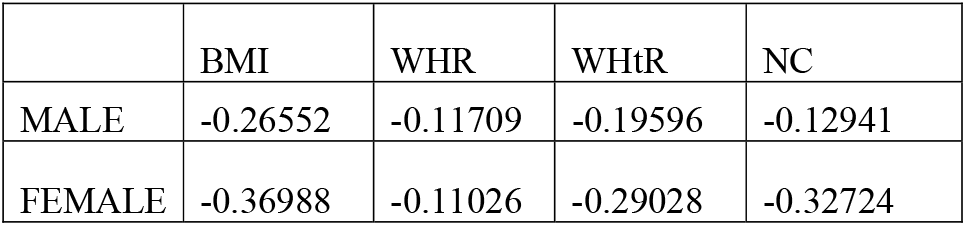
Best predictor of the FI among males and females.

|  | BMI | WHR | WHtR | NC |
| --- | --- | --- | --- | --- |
| MALE | -0.26552 | -0.11709 | -0.19596 | -0.12941 |
| FEMALE | -0.36988 | -0.11026 | -0.29028 | -0.32724 |

## RESULTS

From the above analysis, it was found that all the body compositions measured had a negative correlation with fitness index among both genders which states that increase in any one of the body compositions leads to decrease in fitness index or vice-versa. Body Mass Index, Waist-to-Height Ratio, Neck Circumference are more negatively correlated with Fitness Index in females; and in males Waist-to-Hip Ratio is more negatively correlated with Fitness Index. Also Body Mass Index is the best predictor of fitness index among both the genders and the fitness index level of boys were more than girls.

## DISCUSSION

After a randomized selection, 100 students i.e. 50 males and 50 females from PEWS Group of Institutions fulfilling the inclusion criteria were enrolled in the study. All the subjects did complete the Harvard step test according to protocol and none of them were excluded from the study.

Body compositions i.e. Body Mass Index, Waist-to-hip ratio, Waist-to-height ratio, Neck circumference were measured and were correlated with the Fitness index determined using Harvard step test. Statistical correlation was done using Karl Pearson’s Coefficient of Correlation.

It was found that all the body compositions measured had a negative correlation with fitness index among both genders which states that increase in any one of the body compositions leads to decrease in fitness index or vice-versa. Body Mass Index, Waist-to-Height Ratio, Neck Circumference are more negatively correlated with Fitness Index in females; and in males Waist-to-Hip Ratio is more negatively correlated with Fitness Index. Also Body Mass Index is the best predictor of fitness index among both the genders and the fitness index level of boys were more than girls.

Here most of the body compositions of females have greater negative correlation than the males, which determines that a minor or major increase in those body composition will decrease their fitness index more rapidly or vice-versa. But as males have less negative correlation so an increase or decrease in body compositions will not have much impact on their fitness index. So, the males have better fitness index than the females. Thus, the females are more prone to develop health related issues.

Rohallah Arabmokhtari et al.(2018), conducted a study among students of age group 22-36 years, where they found Body Mass Index and Waist-to-hip ratio is negatively correlated with cardio-respiratory fitness in both the genders and also the females were found to be less fit than males. And subsequently it was found that lower cardio-respiratory fitness students were more likely to be overweight and obese than those with high cardio-respiratory fitness^[12]^. Similarly in an another study conducted by Redzal Abu Hanifah et al.(2014) amongst 13 years old Malaysian students to assess the fitness level and body composition indices which concluded that Body Mass Index, Waist-to-height ratio and waist circumference were negatively correlated with fitness index and the girls to boys fitness ratio was 1:10^[13]^. In the present study too it is found that males are more fitter than the females by assessing the mean values of the fitness index.

In an another study Jing-ya Zhou et al.(2013) on 20-85 years age group to investigate whether neck circumference independently contributes to the prediction of cardio-metabolic risks and thereby found that neck circumference is positively related withcardio-metabolic risk factors amongst both genders; and neck circumference was a more reliable indices to indicate central obesity^[14]^. Similarly Ignacio Ara et al.(2012)conducted a study among 7-12 years children and predicted Body Mass Index and fatmass had strong relationship with cardio-respiratory fitness^[15]^.

Hormones drive the deposition of fat around the pelvis, buttocks, thighs, neck of women and the bellies of men. Genetically research says that as the females have two copies of X chromosome variant, so they tend to be shorter in height than males. Throughout most of their lives females have a higher percentage of body fat than males^[16]^. Most of the females suffer from poor body image which can be lack of willingness to participate in physical activity, more preference for shopping, going to movies, social media and other activities which take up their time. But most of the males nowadays are fitness freak; they prefer to spend their leisure time in gyms ^[17]^.

Thus, from the study it becomes clear that body compositions are correlated with fitness level. Also due to biological development and willingness of participation in physical activities, the fitness levels are more in males than females.

## Data Availability

All data provided is valid and reliable. All data hard and soft copies are available.

## ACKNOWLEDGEMENT

First and foremost, I thank my **ALMIGHTY** without whose blessings and divine wisdom I won’t be able to complete the study in time. I would like to express my warm regards to my parents and friends who always encouraged me to pursue my dreams and have provided untiring support to achieve it.

I am deeply indebted to my teachers **Dr. Dony Kotoky Sarma (PT)**,**Dr. Barnali Bhattacharjee (PT), Dr. Ujwal Bhattacharya (PT)** without whose valuable guidance and suggestions a study of such a magnitude would not have been materialized.

My sincere thanks to **Chumchum Doloi Sarma** ma’am and **Himadri Chutia** ma’am for helping and guiding me in the statistical analysis of my study.

I would like to thanks all the subjects who participated in the study without whom the study would not have been completed.

## REFERENCES

1. Virali Bhansali, Zainab Bharmal. Assessing the cardiovascular fitness of healthy young individuals using Harvard Step Test. School of physiotherapy R.K University, July 2015, Rajkot.

2. Frownfelter, D., & Dean, E. (2012). Cardiovascular and Pulmonary Physical Therapy. In Elsevier.

3. Magalhães, E. I. da S., Sant’ana, L. F. da R., Priore, S. E., & Franceschini, S. do C. C. (2014). Waist circumference, waist/height ratio, and neck circumference as parameters of central obesity assessment in children. In Revista Paulista de Pediatria (Vol. 32, Issue 3). https://doi.org/10.1590/1984-0462201432320

4. Liberato, S. C., Maple-Brown, L., Bressan, J., & Hills, A. P. (2013). The relationships between body composition and cardiovascular risk factors in young Australian men. Nutrition Journal, 12(1). https://doi.org/10.1186/1475-2891-12-108

5. Brouha, L. (1943). The step test: A simple method of measuring physical fitness for muscular work in young men. Research Quarterly of the American Association for Health, Physical Education and Recreation, 14(1). https://doi.org/10.1080/10671188.1943.10621204

6. ^ Top End Sports, Description of the Harvard Step Test.

7. World Health Organization, STEPwise approach to surveillance (STEPS). Retrived March 21, 2012.

8. Ashwell, M., Gunn, P., & Gibson, S. (2012). Waist-to-height ratio is a better screening tool than waist circumference and BMI for adult cardiometabolic risk factors: Systematic review and meta-analysis. In Obesity Reviews (Vol. 13, Issue 3). https://doi.org/10.1111/j.1467-789X.2011.00952.x

9. Schneider, H. J., Friedrich, N., Klotsche, J., Pieper, L., Nauck, M., John, U., Dörr, M., Felix, S., Lehnert, H., Pittrow, D., Silber, S., Völzke, H., Stalla, G. K., Wallaschofski, H., & Wittchen, H. U. (2010). The predictive value of different measures of obesity for incident cardiovascular events and mortality. Journal of Clinical Endocrinology and Metabolism, 95(4). https://doi.org/10.1210/jc.2009-1584

10. Luo Y, Ma X, Shen Y, et al. Neck circumference as an effective measure for identifying cardiometabolic syndrome: a comparison with waist circumference. Endocrine, 31 Oct 2016; Vol 55, Issue 3; 822–830.

11. Patil, C. R., Deshmukh, J. S., Lalwani, C. R., & Bagde, H. (2018). Neck circumference: an alternative tool for screening cardiovascular risk among adults. Journal of Clinical and Diagnostic Research, 12(3). https://doi.org/10.7860/JCDR/2018/30555.11235

12. Arabmokhtari, R., Khazani, A., Bayati, M., Barmaki, S., & Fallah, E. (2018). Relationship between Body Composition and Cardiorespiratory Fitness in Students at Postgraduate Level. Zahedan Journal of Research in Medical Sciences, 20(2). https://doi.org/10.5812/zjrms.12109

13. Hanifah, R. A., Majid, H. A., Jalaludin, M. Y., Al-Sadat, N., Murray, L. J., Cantwell, M., Su, T. T., & Nahar, A. M. (2014). Fitness level and body composition indices: Cross-sectional study among Malaysian adolescent. BMC Public Health, 14. https://doi.org/10.1186/1471-2458-14-S3-S5

14. Zhou, J. ya, Ge, H., Zhu, M. fan, Wang, L. jun, Chen, L., Tan, Y. zong, Chen, Y. ming, & Zhu, H. lian. (2013). Neck circumference as an independent predictive contributor to cardio-metabolic syndrome. Cardiovascular Diabetology, 12(1). https://doi.org/10.1186/1475-2840-12-76

15. Ara, I., Moreno, L. A., Leiva, M. T., Gutin, B., & Casajús, J. A. (2007). Adiposity, physical activity, and physical fitness among children from Aragón, Spain. Obesity, 15(8). https://doi.org/10.1038/oby.2007.228

16. University of Helsinki researchers. Why men are typically taller than women. 6 March 2017.

17. Telford, R. M., Telford, R. D., Olive, L. S., Cochrane, T., & Davey, R. (2016). Why are girls less physically active than boys? Findings from the LOOK longitudinal study. PLoS ONE, 11(3). https://doi.org/10.1371/journal.pone.0150041

